# Clinical prediction models to diagnose neonatal sepsis in low-income and middle-income countries: a scoping review

**DOI:** 10.1101/2024.09.05.24313133

**Authors:** Samuel R Neal, Sarah Sturrock, David Musorowegomo, Hannah Gannon, Michele Zaman, Mario Cortina-Borja, Kirsty Le Doare, Michelle Heys, Gwen Chimhini, Felicity Fitzgerald

## Abstract

Neonatal sepsis causes significant morbidity and mortality worldwide but is difficult to diagnose clinically. Clinical prediction models (CPMs) could improve diagnostic accuracy. Neonates in low-income and middle-income countries are disproportionately affected by sepsis, yet no review has comprehensively synthesised CPMs validated in this setting. We performed a scoping review of CPMs for neonatal sepsis diagnosis validated in low-income and middle-income countries. From 4598 unique records, we included 82 studies validating 44 distinct models. Most studies were set in neonatal intensive or special care units in middle-income countries and included neonates already suspected of sepsis. Three quarters of models were only validated in one study. Our review highlights several literature gaps, particularly a paucity of studies validating models in low-income countries and the WHO African region, and models for the general neonatal population.

Furthermore, heterogeneity in study populations, definitions of sepsis and reporting of models may hinder progress in this field.

## INTRODUCTION

Neonatal sepsis is a clinical syndrome caused by severe systemic infection in the first month of life.^1^ It is a leading cause of global neonatal mortality and disproportionately affects neonates in low-income and middle-income countries (LMICs).^2^

Conventionally, neonatal sepsis is divided into early-onset sepsis (EOS) usually due to vertical transmission of pathogens from the maternal genitourinary tract and occurring in the first 48-72 hours of life, and late-onset sepsis (LOS) usually due to pathogens acquired from the home or hospital environment and occurring after 48-72 hours.^3^ For infections caused by group B streptococcus (GBS), EOS is often considered to occur up to the seventh day of life.^3^ However, there is increasing recognition that this conventional classification of neonatal sepsis is misplaced in LMICs where neonates may be exposed from birth to organisms traditionally associated with LOS due to high rates of healthcare-associated infections.^4^

Neonatal sepsis is difficult to diagnose clinically due to non-specific signs and symptoms. Identifying a pathogenic organism from a normally sterile site (e.g. blood or cerebrospinal fluid) remains the gold standard method for diagnosis.^3^ Nevertheless, *‘*clinical’ or ‘culture-negative’ sepsis – where a sterile culture is obtained from a neonate with signs and symptoms of sepsis – is a recognised entity.^5^ When sepsis is suspected, a fine balance must be struck between the risk of failing to treat a true invasive infection and the risks of unnecessary antimicrobial use, which can contribute to antimicrobial resistance,^4^ and adverse neonatal outcomes.^6^ Additionally, national guidelines advise starting antimicrobial therapy within one hour of suspecting sepsis to maximise the chance of survival, leaving little time for clinicians to make a diagnosis.^7^

Clinical prediction models (CPMs) are tools that combine characteristics to estimate the probability of a diagnosis or prognostic outcome.^8^ Models exist to diagnose neonatal sepsis in both high-income countries and LMICs based on various clinical features, risk factors and/or laboratory tests. These CPMs aim to improve diagnostic accuracy and therefore rationalise antibiotic use. The benefits of early recognition and treatment could be significant for neonates in low-resource settings where specialist care is limited.^9^ Furthermore, reducing the growing threat of antimicrobial resistance is even more crucial where access to sufficiently broad-spectrum antimicrobials may be unaffordable for patients.^10^

While several existing reviews examine CPMs to diagnose neonatal sepsis,^11–14^ there has been no comprehensive synthesis of models validated in LMICs. Therefore, the aim of this scoping review was to map the literature of CPMs to diagnose neonatal sepsis in LMICs.

Specific objectives were:

1. To provide an overview of existing CPMs to diagnose neonatal sepsis in LMICs
2. To determine the evidence underlying the use of CPMs to diagnose neonatal sepsis in LMICs
3. To compare the performance of CPMs using different approaches to risk stratification or different target populations
4. To identify unanswered research questions surrounding CPMs to diagnose neonatal sepsis in LMICs, which may guide future primary research or systematic reviews

## METHODS

We conducted this review according to an *a priori* published protocol,^15^ developed with reference to the scoping review guidelines provided by the Joanna Briggs Institute.^16^ We report methods and results in accordance with the Preferred Reporting Items for Systematic reviews and Meta-Analyses extension for Scoping Reviews (see supplementary appendix).^17^

### Search strategy

Eligibility criteria are shown in Table 1. After reviewing the extent and breadth of the literature from our initial searches, we narrowed the scope of our original protocol to focus specifically on studies that validate a CPM to diagnose neonatal sepsis in a LMIC, as defined by the World Bank in 2020.^18^ We searched six electronic databases from their inception: Ovid MEDLINE, Ovid Embase, Scopus, Web of Science Core Collection, Global Index Medicus, and the Cochrane Library. Searches were initially performed on 20 December 2019 and updated on 5 September 2022 and 16 June 2024.

**Table 1.** Eligibility criteria.

|  |  |
| --- | --- |
| **Population** | • Neonates aged $\leq 30$ days of life, or hospitalised in a neonatal unit, being evaluated for neonatal sepsis.<br>• The definition of neonatal sepsis was left to the discretion of individual studies.<br>• Studies examining a range of patient ages were included, provided that sufficient data were available to examine model performance in neonates specifically. |
| **Concept** | • Studies that develop or validate a CPM to diagnose neonatal sepsis in a LMIC, as defined by the World Bank in 2020.<br>• Studies were considered to have validated a CPM if they reported any of: C-statistic denoting AUC, sensitivity, specificity, predictive values or likelihood ratios.<br>• Studies evaluating prognostic models (e.g. to predict mortality or morbidity) were excluded. |
| **Context** | • Studies from any healthcare setting, including neonatal units, emergency departments, outpatient settings, or community settings.<br>• Studies were only included if the model they refer to has been internally and/or externally validated in a LMIC, regardless of the country where the model was initially developed. |
| **Type of studies** | • Randomised and quasi-randomised controlled trials, cohort studies, cross-sectional studies, case-control studies and clinical guidelines.<br>• Also, letters, comments and conference proceedings (providing sufficient details were provided for data extraction).<br>• Systematic reviews, meta-analyses and editorials were excluded, but were used to identify relevant primary literature.<br>• No time period restrictions, but only studies published in English or Spanish were considered, to reflect the languages spoken by the review team. |

AUC = area under the receiver operating characteristic curve; CPM = clinical prediction model; LMIC = low-income and middle-income country.

Search terms were chosen to capture the three domains of the research question (‘neonate’, ‘sepsis’, and ‘clinical prediction model’) through collaboration with a child health specialist librarian. The search strategy was developed for Ovid MEDLINE and adapted for each database (see supplementary appendix). Additional studies were identified by citation analysis and by hand searching the reference lists of included studies.

### Record screening

We imported identified records into EndNote 21 for deduplication.^19^ Unique records were then uploaded to the Rayyan application for screening by two independent reviewers (DM, HG, MZ, SRN or SS).^20^ Titles and abstracts were first examined against the eligibility criteria to determine if each record was potentially eligible for inclusion. Next, full texts of potentially eligible studies were obtained and reviewed to confirm eligibility. Authors were contacted to request full texts where these could not be found online. Conflicts were resolved by discussion amongst the review team.

### Data extraction and synthesis

Data extraction was performed independently by two reviewers for the initial searches (SRN and SS) and by one reviewer for each updated search (SRN or SS). We extracted data on study, participant and model characteristics, and model performance using a pre-piloted data extraction form (see supplementary appendix). Data items were chosen based on the Transparent Reporting of a multivariable prediction model for Individual Prognosis Or Diagnosis (TRIPOD) statement.^21^ We summarised results by narrative synthesis. Data for quantitative outcomes were not pooled in a meta-analysis as this is beyond the scoping review methodology. Where multiple variations of a model were presented in the same study (e.g. different combinations of predictors presented during model specification), or model performance was presented at multiple classification thresholds, we only included data for the ‘optimal’ or ‘final’ model at a single classification threshold.

## RESULTS

### Searches and included studies

Searches identified 4598 unique records (Figure 1). From these, 82 studies published between 2003 and 2024 were included,^22–103^ and are summarised in Tables 2 and 3. The number of published studies validating a CPM to diagnose neonatal sepsis in LMICs has increased rapidly in recent years (Figure 2). Studies were conducted in 22 individual countries (Figure 3 and Table 4), with the greatest number of studies conducted in the World Health Organization (WHO) South-East Asian Region (*n=*48, 59%), particularly in India (*n=*37, 45%). The fewest studies were conducted in the WHO African Region (*n=*4, 5%). Regarding economic status, 51 studies were conducted exclusively in lower middle-income countries (62%) and 30 exclusively in upper middle-income countries (37%).

**Figure 1.**
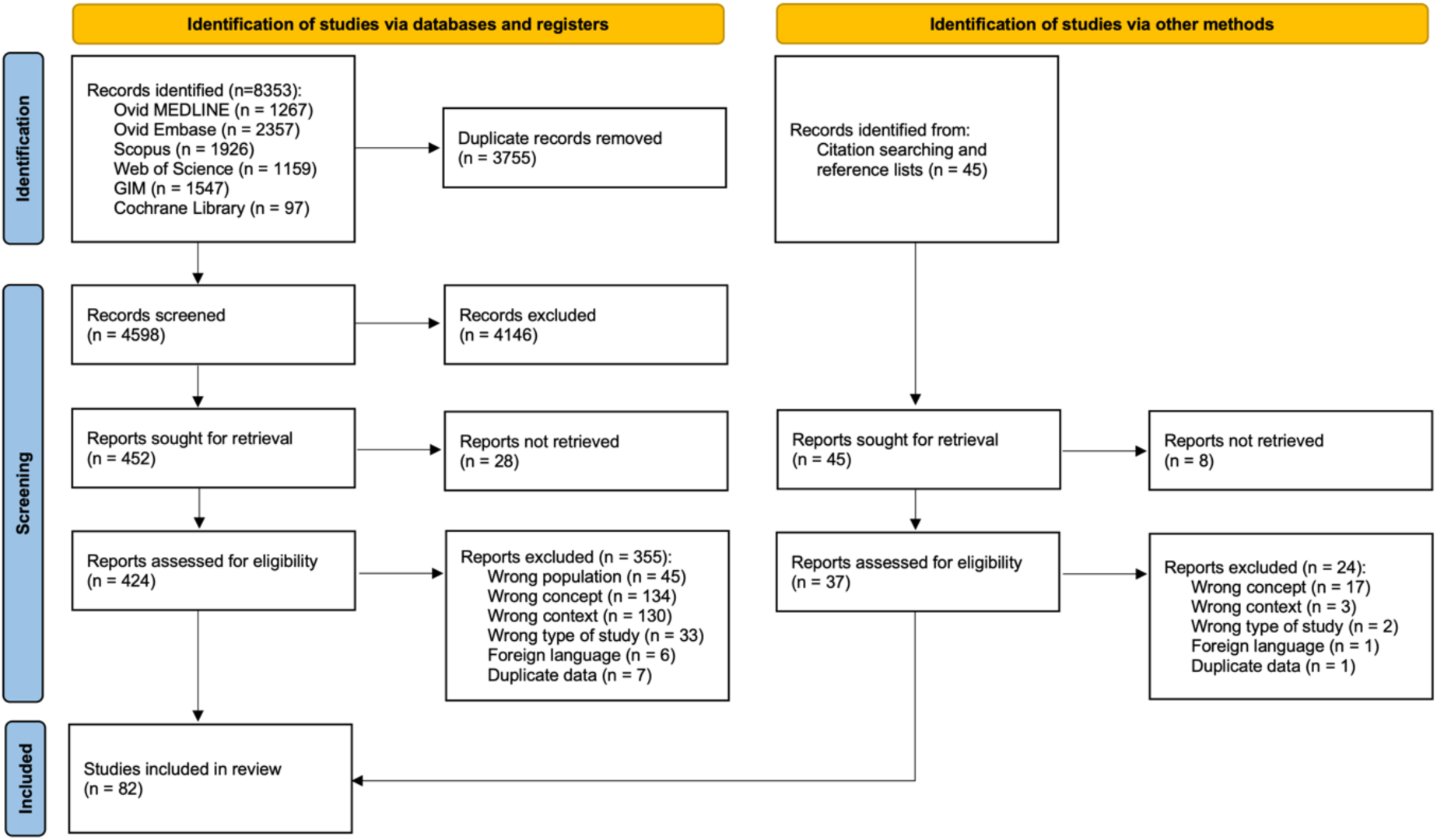
PRISMA flowchart of study selection. GIM = Global Index Medicus. Adapted from Page MJ, McKenzie JE, Bossuyt PM, Boutron I, Hoffmann TC, Mulrow CD, et al. The PRISMA 2020 statement: an updated guideline for reporting systematic reviews. BMJ 2021;372:n71. doi: 10.1136/bmj.n71.

**Figure 2.**
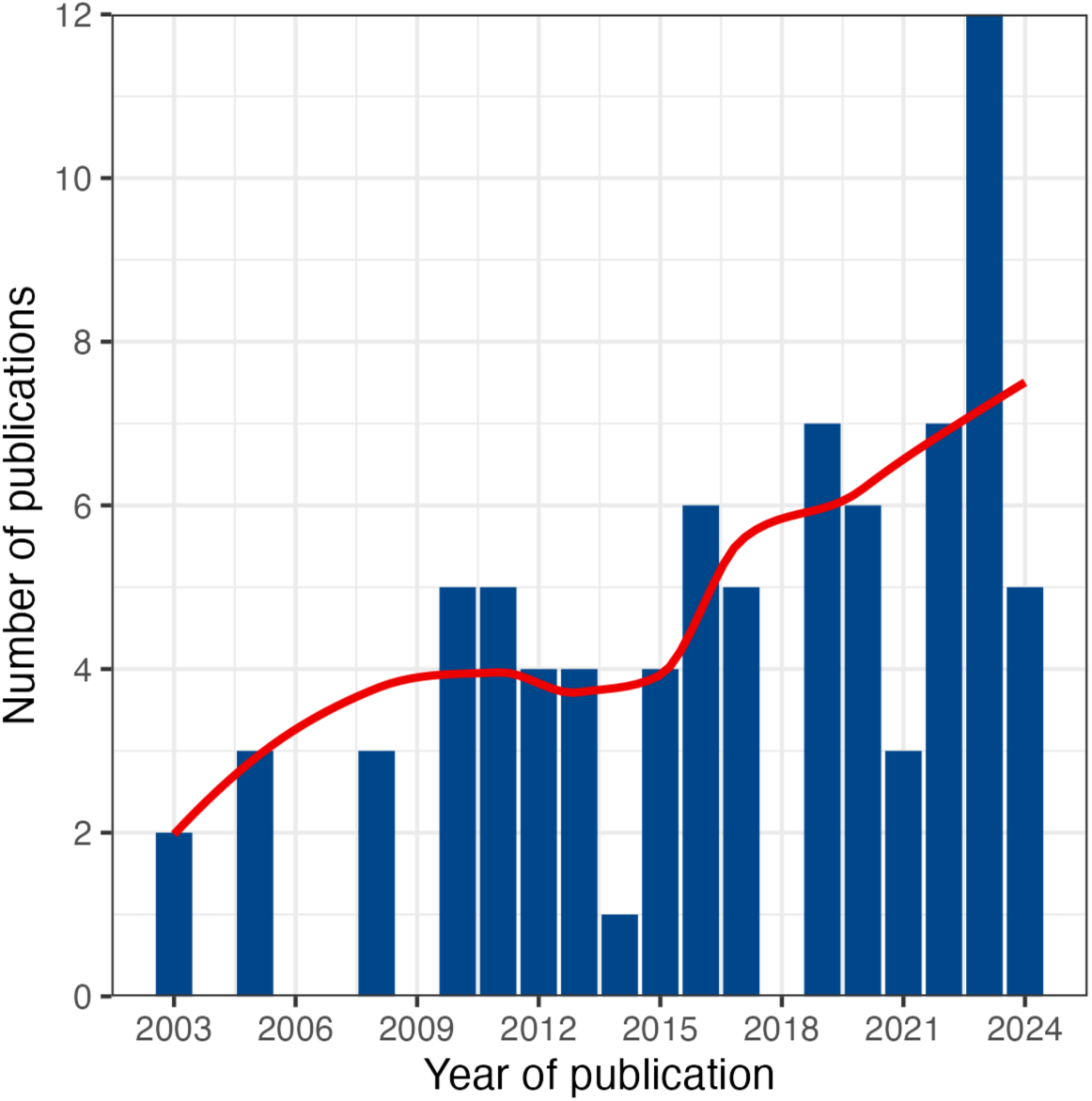
Number of studies included in our review by year of publication. Red line represents a local regression (LOESS) model fitted on the yearly publication counts.

**Figure 3.**
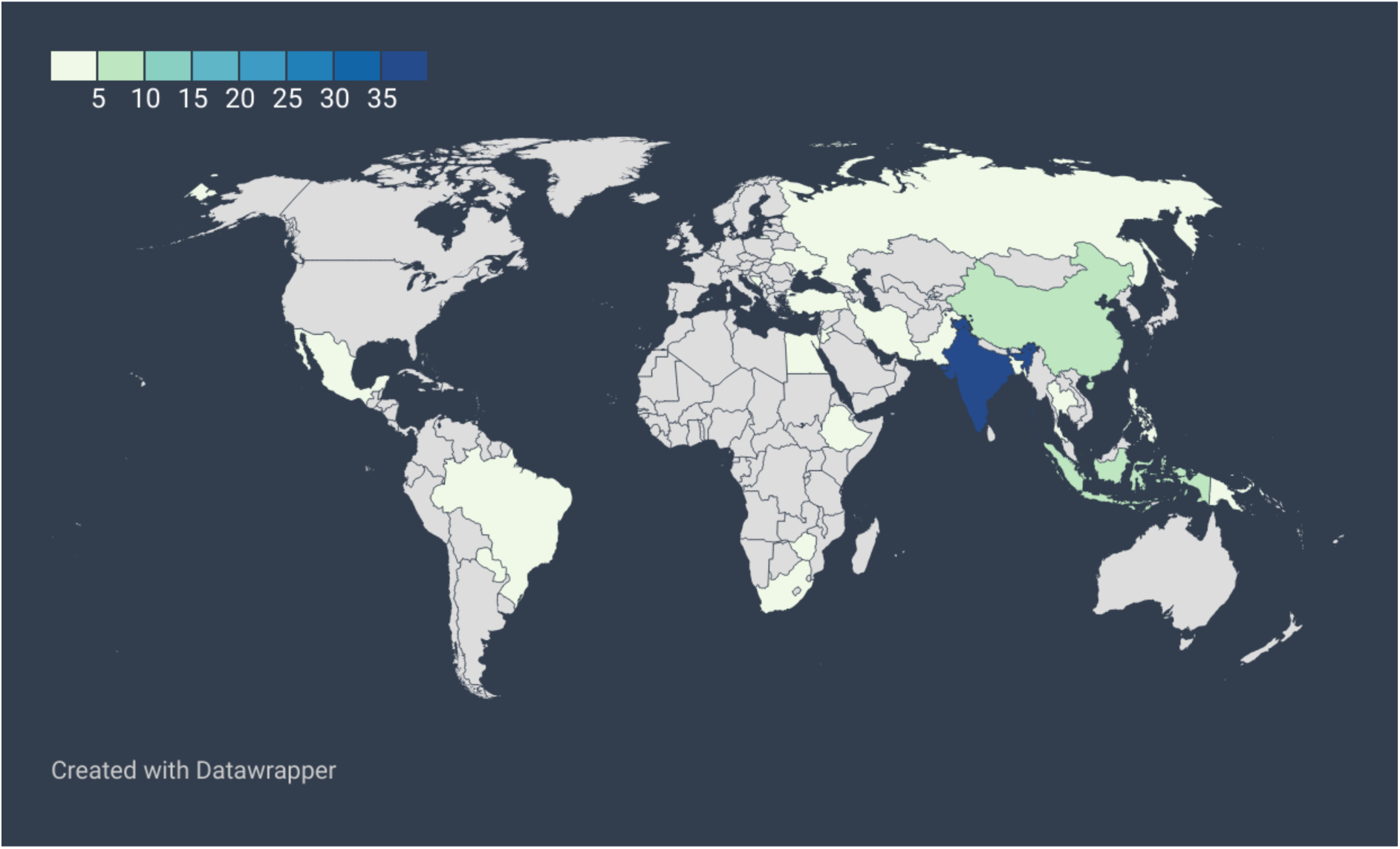
Choropleth map of number of included studies by country. Created with DataWrapper: Lorenz, Aisch, and Kokkelink, ‘Datawrapper: Create Charts and Maps’.

**Table 2.**
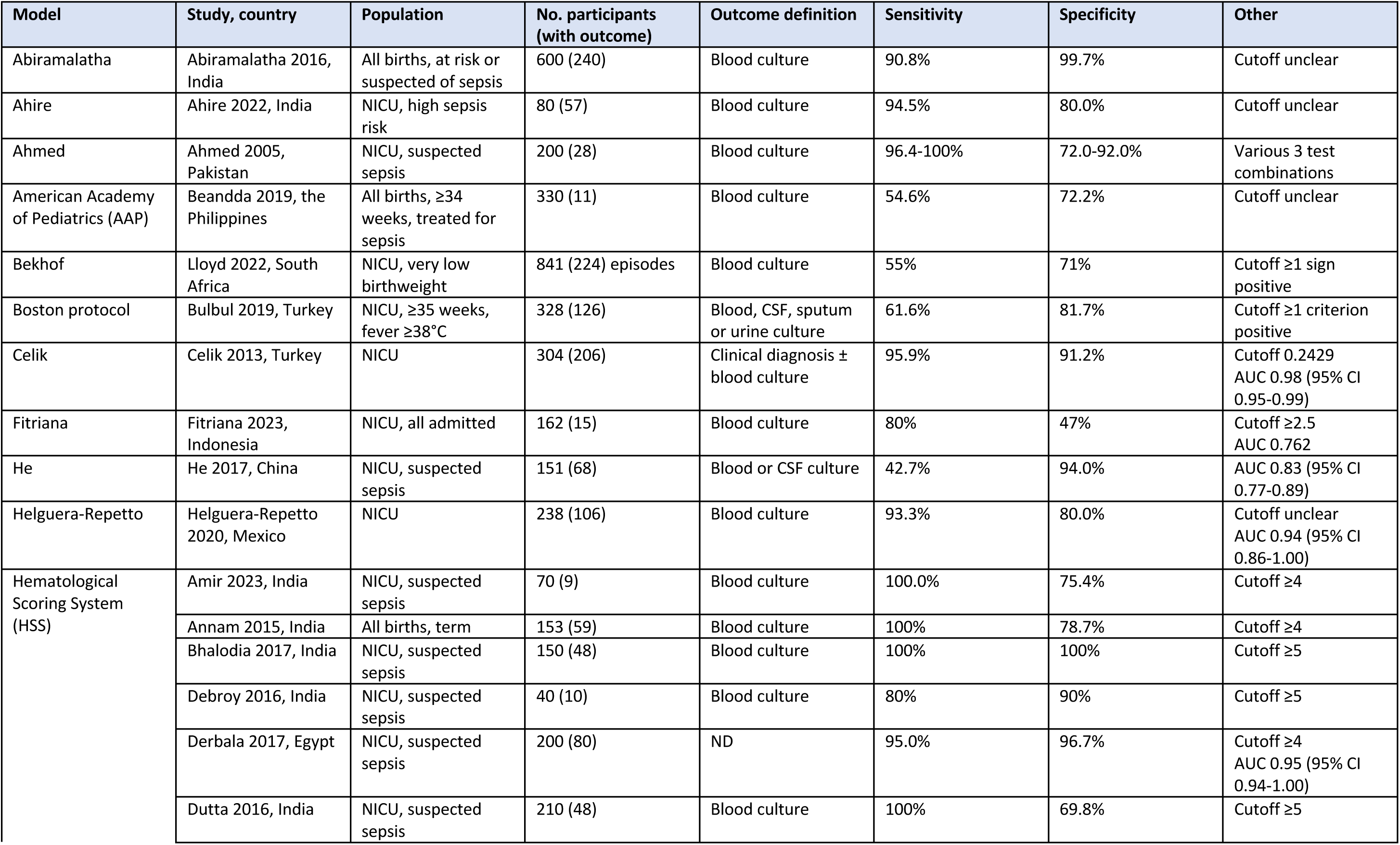
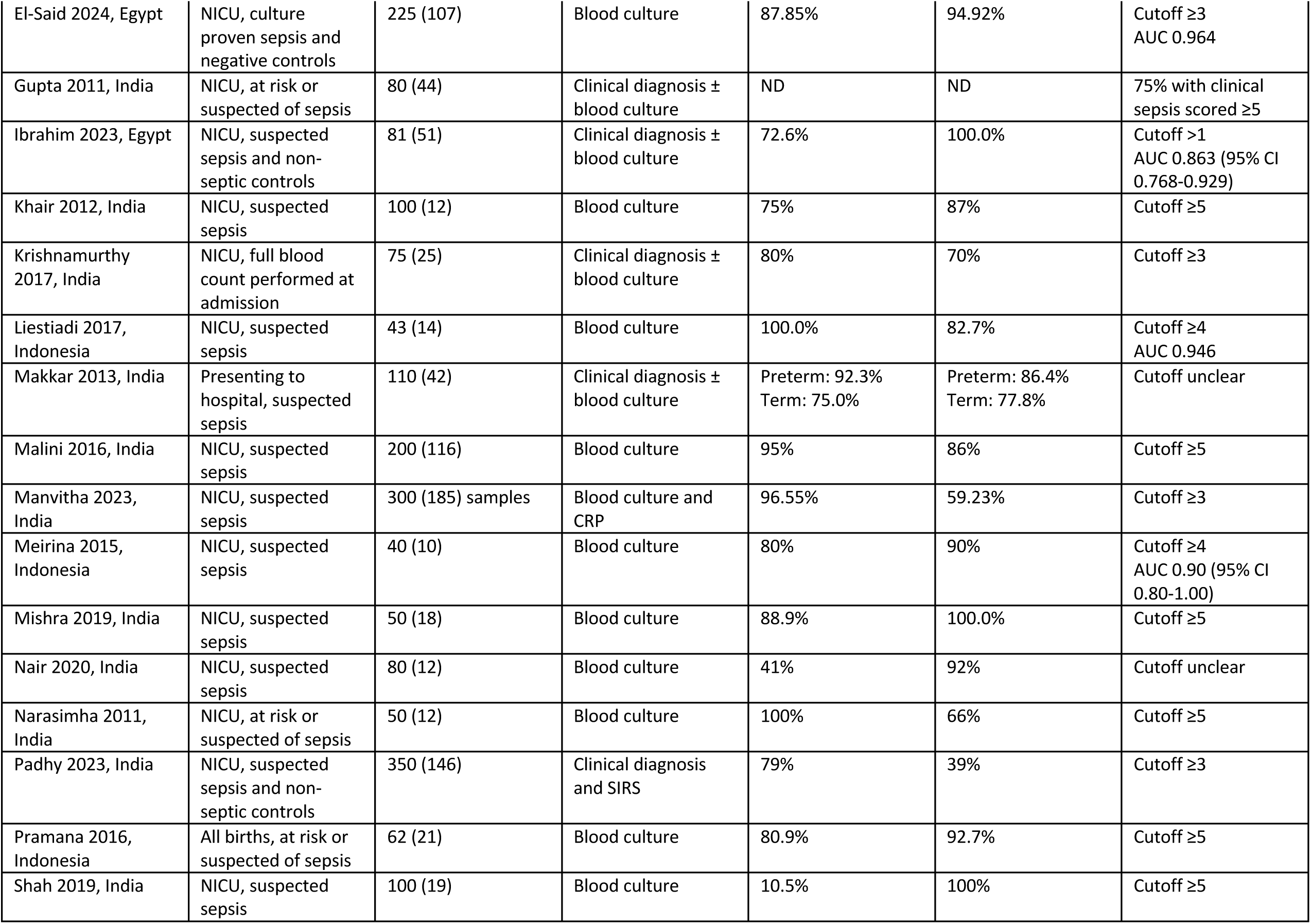
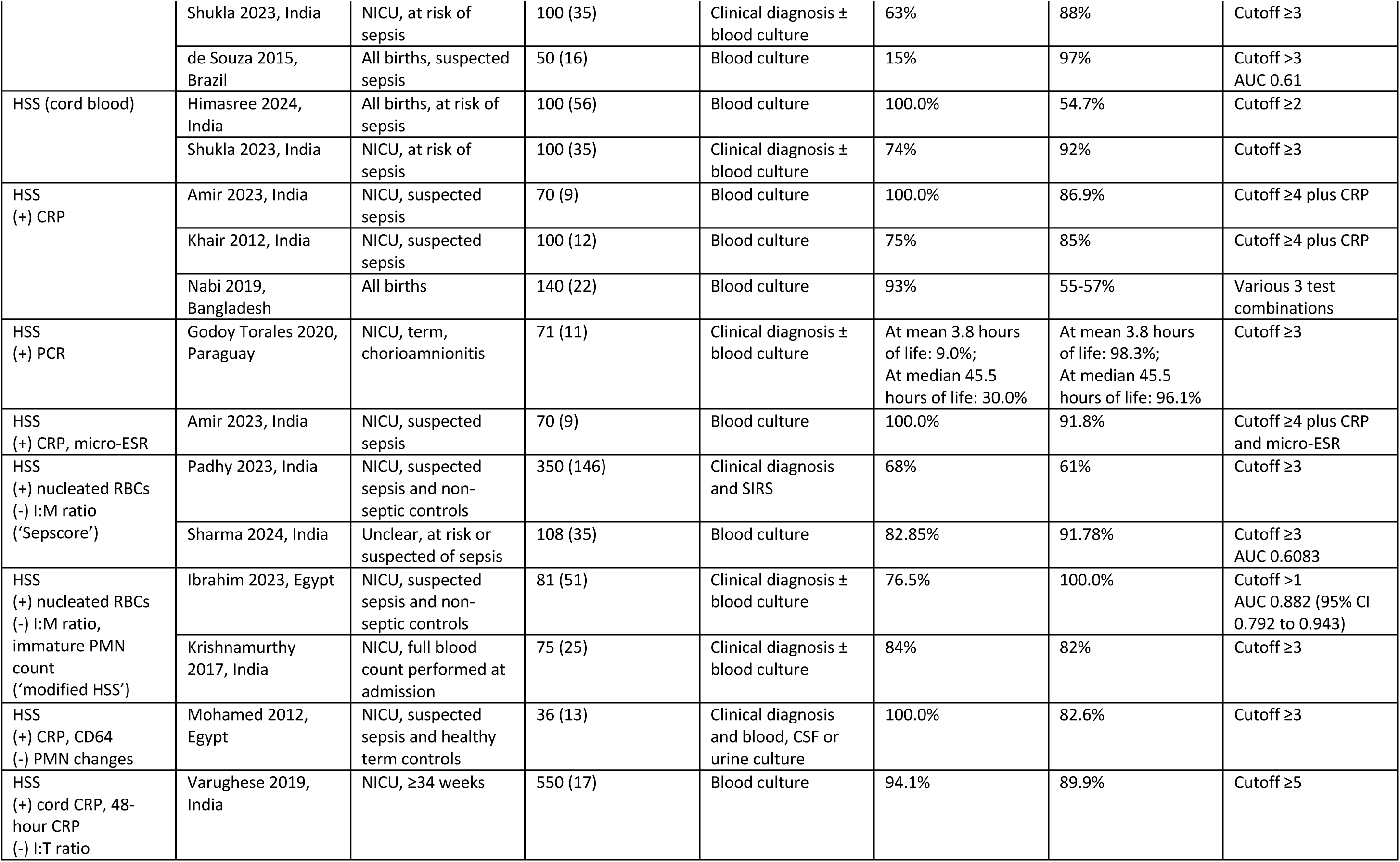
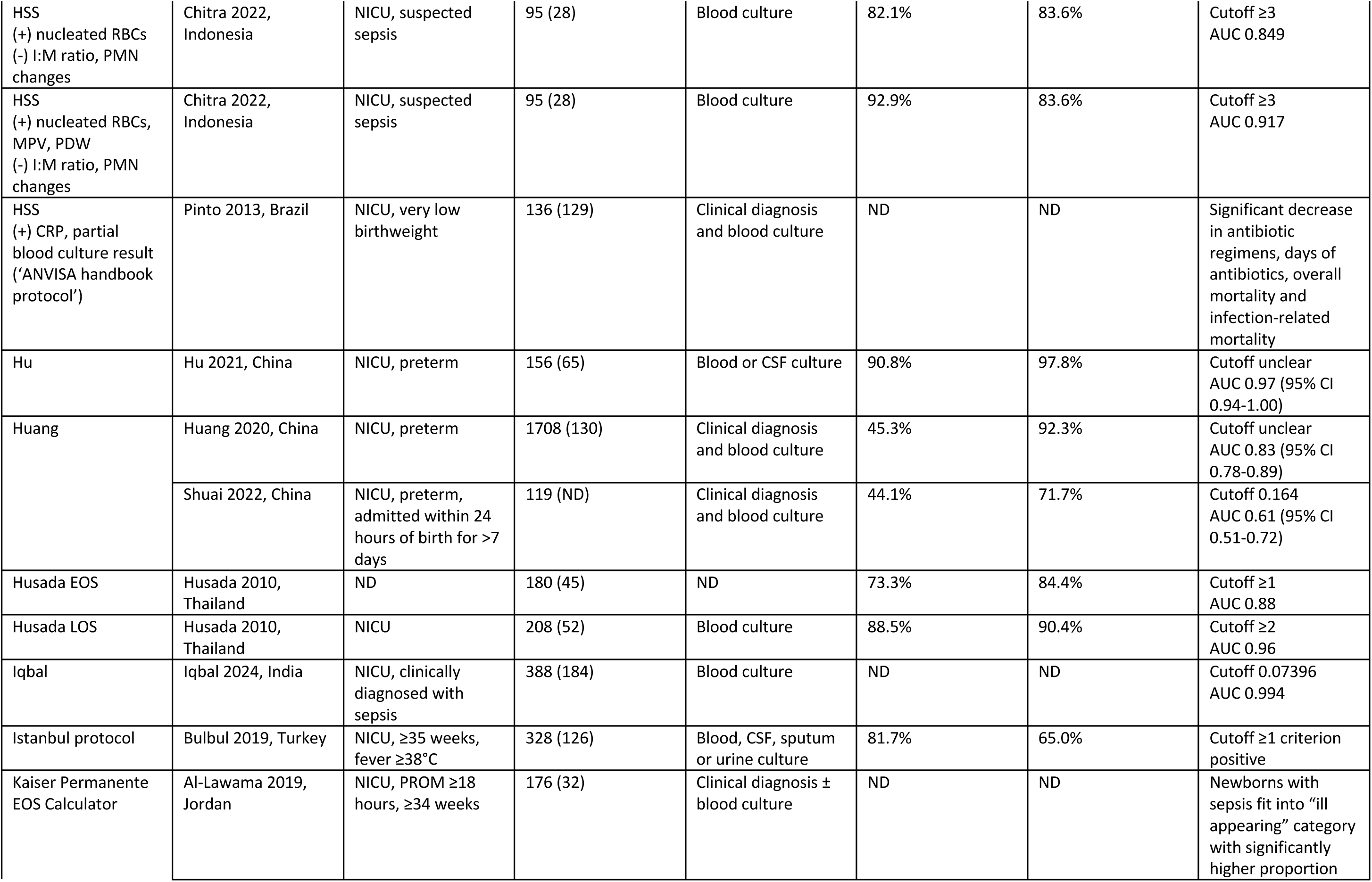
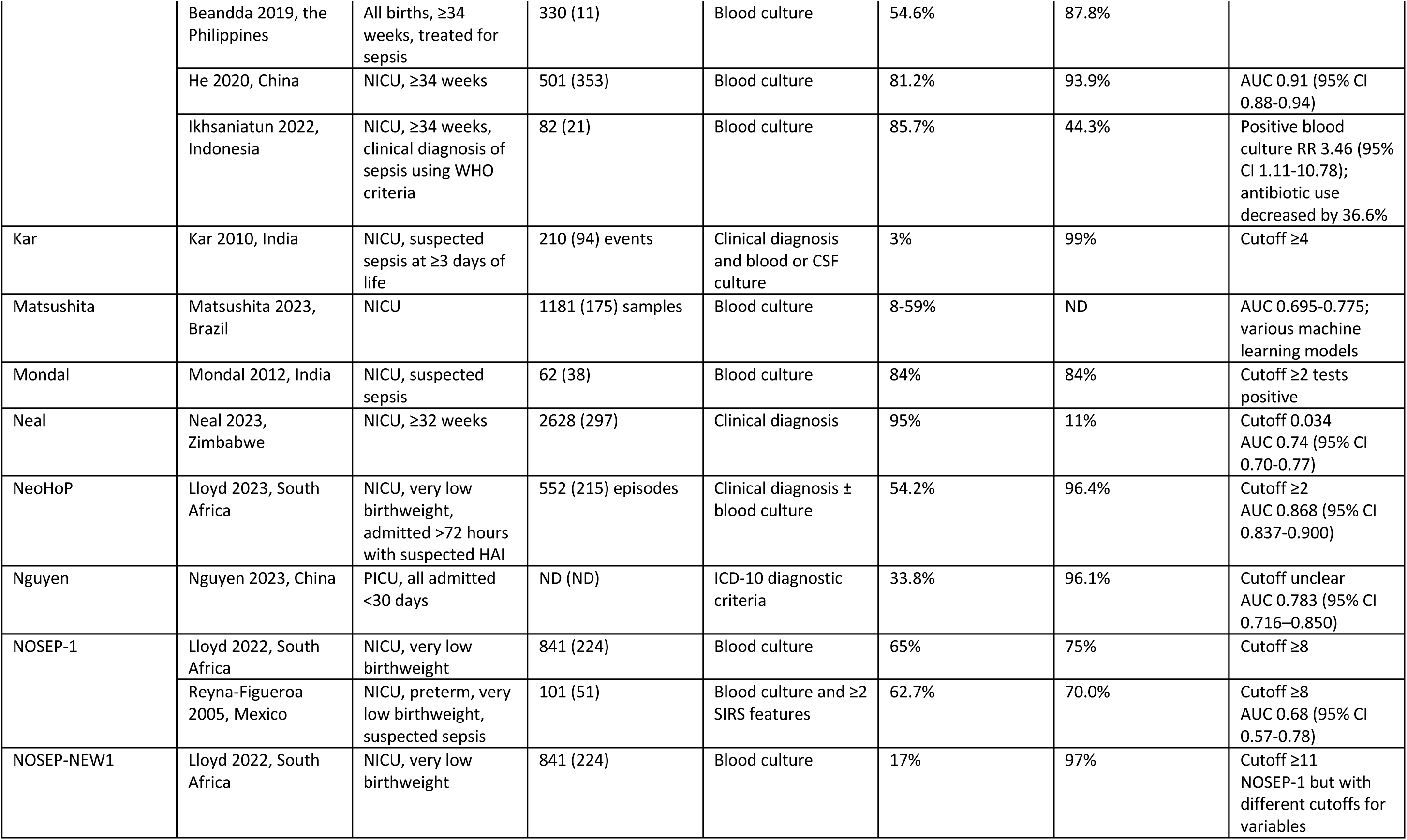
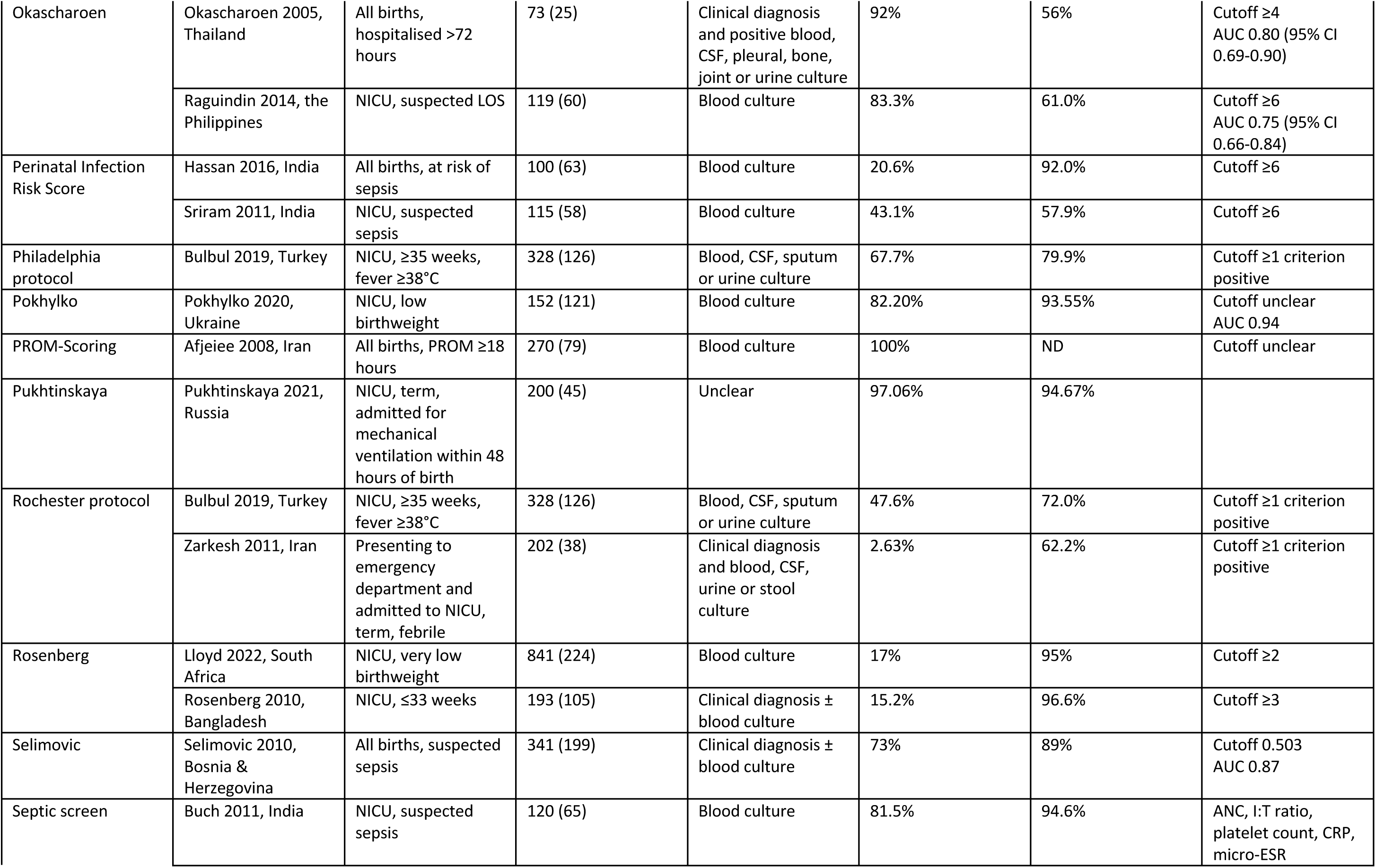
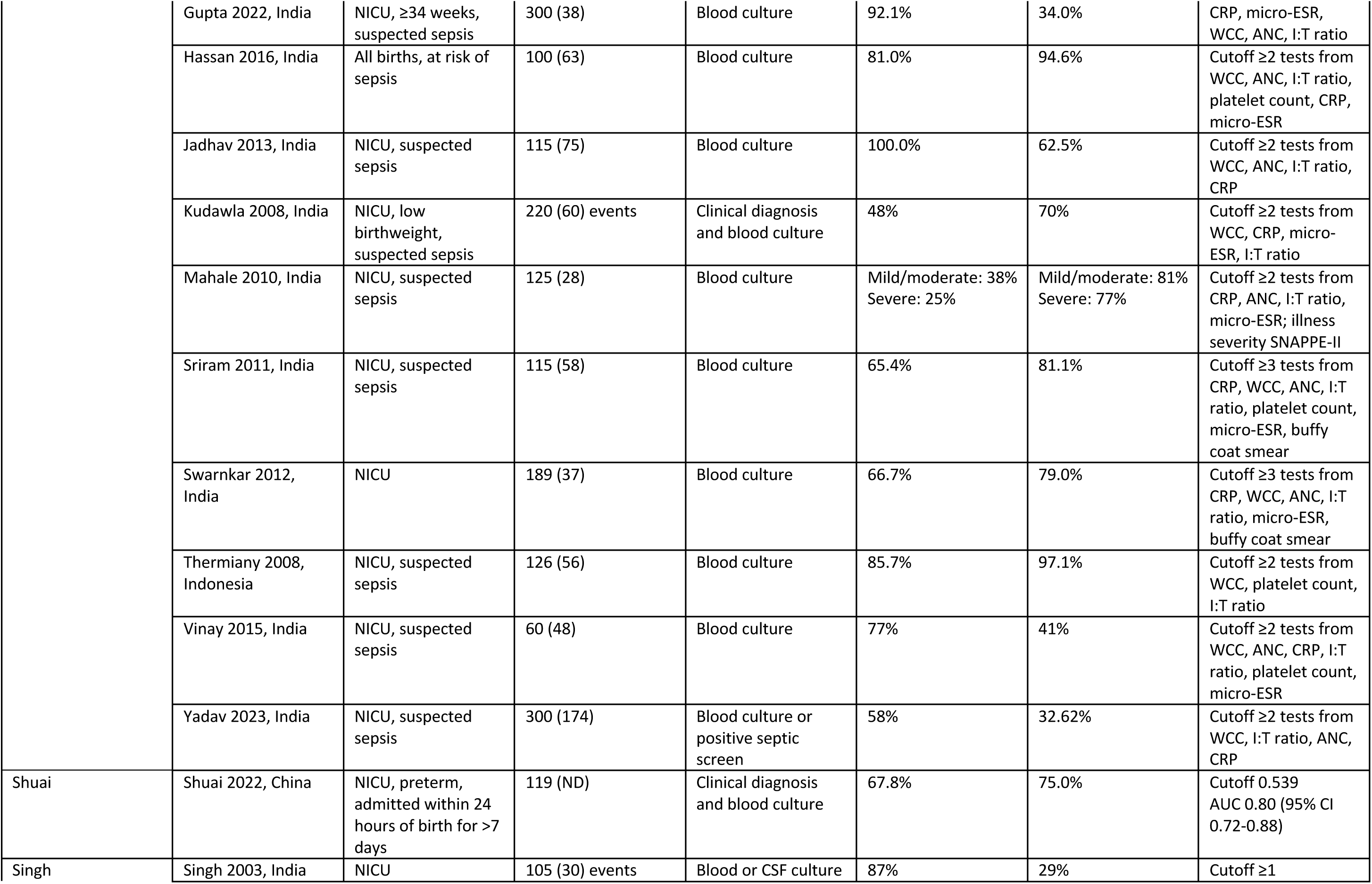
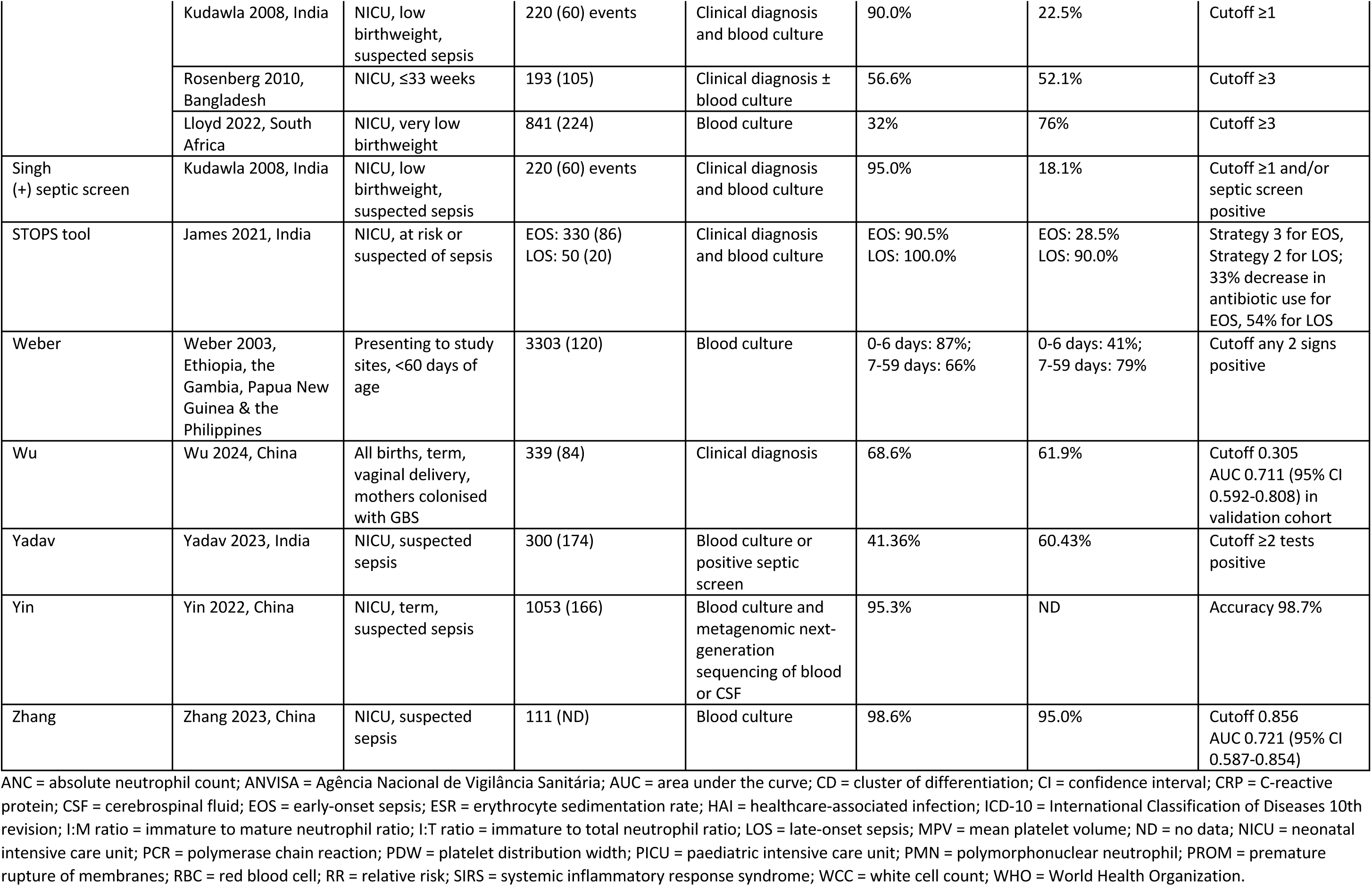
Summary of included studies and model performance.

**Table 3.** Summary of model characteristics.

| Model (derivation study) | Country of derivation cohort | Outcome | Modelling methods | Predictors in final model |  |  |
| --- | --- | --- | --- | --- | --- | --- |
|  |  |  |  | Clinical features | Risk factors | Laboratory tests |
| Abiramalatha (Abiramalatha 2016) | India | All sepsis | Scoring system, ROC curves to determine cutoff values |  |  | WCC, platelet count, lower median angle light scatter, mean neutrophil volume |
| Ahire (Ahire 2022) | India | All sepsis | Scoring system |  |  | CRP, WCC, platelet count, neutrophil count |
| Ahmed (Ahmed 2005) | Pakistan | All sepsis | Scoring system, literature and clinical knowledge |  |  | CRP, neutrophil count, platelet count, gastric aspirate cytology, cytoplasmic vacuolation (3 test combinations) |
| American Academy of Pediatrics (AAP) (Puopolo 2018) | USA | EOS | Criteria-based without specific scoring | Ill appearance | Chorioamnionitis |  |
| Bekhof (Bekhof 2013) | The Netherlands | LOS | Nomogram from logistic regression, backward elimination | Increased respiratory support, capillary refill time, grey skin | Central venous catheter |  |
| Boston protocol (Baskin 1992) | USA | Serious bacterial infection | Criteria-based without specific scoring | Fever $\geq 38^{\circ}\text{C}$, appearance, dehydration, focal signs of infection | Recent immunisation, recent antibiotics | WCC, CSF analysis, urinalysis, CXR |
| Celik (Celik 2013) | Turkey | All sepsis | Markov state models, modified stepwise selection |  |  | Mean neutrophil volume, volume distribution width, interleukin-6, CRP |
| Fitriana (Fitriana 2023) | Indonesia | EOS | Logistic regression |  | Prematurity, PROM, foul liquor, sex |  |
| Hematological Scoring System (Rodwell 1988) | Australia | All sepsis | Scoring system, univariable predictor performance |  |  | WCC, PMN count, immature PMN count, I:T ratio, I:M ratio, platelet count, PMN degenerative changes |
| He (He 2017) | China | EOS | Logistic regression, stepwise selection |  |  | Interleukin-27, procalcitonin, CRP |
| Helguera-Repetto (Helguera-Repetto 2020) | Mexico | All sepsis | Neural network | Fever >37.5°C, hypothermia <35.5°C, tachycardia, tachypnoea, bradycardia, bradypnoea, apnoeas | PROM, chorioamnionitis, maternal age, maternal morbidity, cervicovaginitis, UTI, sex, gestational age, birthweight, foetal morbidity, catheter, mechanical ventilation | WCC, platelet count, neutrophil count, band cells, % bands, I:T ratio |
| Hu (Hu 2021) | China | EOS | Logistic regression, backward elimination | Apgar score |  | CRP, procalcitonin, interleukin-6 |
| Huang (Huang 2020) | China | LOS | Nomogram from logistic regression, backward elimination |  | Birthweight, intubation, umbilical venous catheter duration | Thyroid function |
| Husada EOS (Husada 2010) | Thailand | EOS | Logistic regression | Oxygen requirement, poor feeding | Length of admission pre-sepsis | WCC, platelet count |
| Husada LOS (Husada 2010) | Thailand | LOS | Logistic regression | Oxygen requirement, poor feeding, abnormal heart rate, abnormal temperature |  | WCC, pH |
| Iqbal (Iqbal 2024) | India | All sepsis | Supervised machine learning | Apnoea, tachycardia, bradycardia, desaturation, lethargy, septic shock, meningitis, pneumonia, Apgar score | Prematurity, vaginal delivery, low birthweight, very low birthweight, IUGR, SGA, primigravida, central catheter, peripheral catheter, end-diastolic flow, CPAP, mechanical ventilation, inotropes, corticosteroids, cardiac disease, lung disorder, respiratory distress syndrome, intraventricular haemorrhage, necrotising enterocolitis, fungal infection | WCC, RBC count, platelet count, neutrophil count, CRP, hypocalcaemia, hypoglycaemia, metabolic acidosis |
| Istanbul protocol (Bulbul 2019) | Turkey | Serious bacterial infection | Criteria-based without specific scoring | Fever ≥38°C, appearance, dehydration, focal signs of infection | Perinatal antibiotics, chronic disease, hospitalised longer than mother | CRP, WCC, I:T ratio, urinalysis |
| Kaiser Permanente EOS Calculator (Kuzniewicz 2017) | USA | EOS | Bayesian logistic regression and recursive partitioning | Clinical presentation (well appearing, equivocal, clinical illness) | EOS incidence, gestational age, duration of ROM, highest maternal intrapartum temperature, maternal GBS, intrapartum antibiotics |  |
| Kar (Kar 2010) | India | LOS | Scoring system, univariable positive likelihood ratio | Lethargy, tachycardia, fever >37.5°C, abdominal distention, increased prefeed aspirate, chest retractions, grunting |  |  |
| Matsushita (Matsushita 2023) | Brazil | All sepsis | Supervised machine learning |  |  | Haemoglobin, haematocrit, MCV, MCH, MCHC, WCC, neutrophil %, neutrophil count, neutrophil left shift %, neutrophil left shift count, eosinophil %, eosinophil count, basophil %, basophil count, lymphocyte %, lymphocyte count, monocyte %, monocyte count, platelet count, neutrophil to lymphocyte ratio, monocyte to lymphocyte ratio, platelet to lymphocyte ratio, delta neutrophil index, CRP, lymphocyte to CRP ratio |
| Mondal (Mondal 2012) | India | All sepsis | Scoring system, univariable preselection |  |  | CRP, micro-ESR, I:T ratio, morphological changes in neutrophils |
| Neal (Neal 2023) | Zimbabwe | EOS | Logistic regression | Fever >37.5°C, respiratory rate, activity, chest retractions, grunting | PROM, maternal intrapartum fever, foul smelling liquor |  |
| NeoHoP (Lloyd 2023) | South Africa | LOS (HAI) | Logistic regression, univariable preselection | Capillary refill time, lethargy, abdominal distention | Central venous catheter | CRP |
| Nguyen (Nguyen 2023) | China | All sepsis | Tree augmented naive Bayes | Overall symptoms, gastrointestinal symptoms, central nervous symptoms, skin symptoms, respiratory symptoms, cardiovascular symptoms, infective symptoms, abnormal temperature symptoms, heart rate, respiratory rate, temperature, oxygen saturation, systolic blood pressure, diastolic blood pressure | Age, gender, low birthweight, prematurity, use of vasoactive agents, ICU length of stay, hospital length of stay, in-hospital mortality | WCC, neutrophil count, lymphocyte count, platelet count, partial thromboplastin time, prothrombin time, ESR, glucose, lactate, creatinine, procalcitonin, CRP, positive microbiological test |
| NOSEP-1 and NOSEP-NEW1 (Mahieu 2000, Mahieu 2002) | Belgium | LOS | Logistic regression, univariable preselection and stepwise selection | Fever >38.2°C | Parenteral nutrition for ≥14 days | CRP, platelet count, neutrophil fraction |
| Okascharoen (Okascharoen 2005) | Thailand | LOS | Cox model, univariable preselection and backward elimination | Hypotension, abnormal temperature, respiratory insufficiency | Umbilical venous catheter duration | Band cell fraction, platelet count |
| Perinatal Infection Risk Score (Takkar 1974) | India | All sepsis | ND | Apgar score | Prematurity, low birthweight, PROM, foul smelling liquor, unclean vaginal examination before delivery, duration of labour exceeding 24 hours |  |
| Philadelphia protocol (Baker 1993) | USA | Serious bacterial infection | Criteria-based without specific scoring | Appearance, focal signs of infection |  | WCC, I:T ratio, CSF analysis, urinalysis, stool analysis, CXR |
| Pokhylko (Pokhylko 2020) | Ukraine | EOS | Logistic regression, stepwise selection | Apgar score | Prematurity, PROM, visual changes in placenta, history of abortion | Monocyte count |
| PROM-Scoring (Afjeiee 2008) | Iran | All sepsis | Scoring system | Apgar score | Prematurity, PROM, foetal tachycardia, chorioamnionitis, low birthweight, sex |  |
| Pukhtinskaya (Pukhtinskaya 2021) | Russia | EOS | Decision tree |  |  | CD95, nitric oxide, CD34, CD69, lymphocytes with expression AnnexinV-FITC+PI |
| Rochester protocol (Powell 1990) | USA | Serious bacterial infection | Criteria-based without specific scoring | Fever ≥38°C, appearance, focal signs of infection | Prematurity, perinatal antibiotics, chronic disease, hospitalised longer than mother | WCC, immature PMN count, urinalysis, stool analysis |
| Rosenberg (Rosenberg 2010) | Bangladesh | LOS | Logistic regression, univariable preselection | Pallor, apnoea, lethargy, jaundice, hepatomegaly |  |  |
| Selimovic (Selimovic 2010) | Bosnia & Herzegovina | EOS | Logistic regression, univariable preselection |  |  | WCC, I:T ratio, I:M ratio, CRP |
| Septic screen (generic) | Various | All sepsis | NA |  |  | CRP, micro-ESR, WCC, neutrophil count, I:T ratio, platelet count |
| Shuai (Shuai 2022) | China | LOS | Logistic regression, univariable preselection |  | Dopamine use, PROM, albumin use, maternal age, gender, peripherally inserted central catheter, gestational age, antibiotic use, season of delivery, asphyxia, prenatal glucocorticoids, umbilical venous catheter, birth weight |  |
| Singh (Singh 2003) | India | LOS | Scoring system, univariable positive likelihood ratio | Lethargy, tachycardia, fever >37.5°C, abdominal distention, increased prefeed aspirate, chest retractions, grunting |  |  |
| STOPS tool (James 2021) | India | All sepsis | Scoring system | Fever >37.5°C, hypothermia <35.5°C, tachycardia, bradycardia, tachypnoea, oxygen requirement, respiratory distress, poor feeding, hypoglycaemia, lethargy, irritability, weak cry, spontaneous movements, seizures, temperature of hands and feet, increased ventilatory requirements, apnoea, capillary refill time, arterial hypotension, skin colour |  | Procalcitonin, glucose |
| Weber (Weber 2003) | Ethiopia, the Gambia, Papua New Guinea & the Philippines | All sepsis, meningitis, pneumonia, or hypoxaemia | Logistic regression, univariable preselection | Fever >38°C, reduced feeding, no spontaneous movement, drowsy or unconscious, history of feeding problems, history of change in activity, agitated, capillary refill time, chest wall indrawing, respiratory rate, grunting, cyanosis, seizures, bulging fontanelle |  |  |
| Wu (Wu 2024) | China | All sepsis | Nomogram from logistic regression, univariable preselection |  | Maternal age, gestational diabetes, forceps delivery, umbilical cord winding, gender |  |
| Yadav (Yadav 2023) | India | All sepsis | Scoring system |  |  | WCC, I:T ratio, neutrophil count, CRP, platelet count, mean platelet volume, platelet distribution width |
| Yin (Yin 2022) | China | Invasive bacterial infection | Decision tree from logistic regression, stepwise selection | Ill appearance, abnormal neurological signs | Age at admission | WCC, procalcitonin, CRP, neutrophil % |
| Zhang (Zhang 2023) | China | EOS | Logistic regression, backward elimination | Vomiting, cough | Age, intra-amniotic infection |  |

CD = cluster of differentiation; CPAP = continuous positive airway pressure; CRP = C-reactive protein; CSF = cerebrospinal fluid; CXR = chest x-ray; EOS = early-onset sepsis; ESR = erythrocyte sedimentation rate; GBS = Group B streptococcus; HAI = healthcare-associated infection; ICU = intensive care unit; I:M ratio = immature to mature neutrophil ratio; I:T ratio = immature to total neutrophil ratio; IUGR = intrauterine growth restriction; LOS = late-onset sepsis; MCH = mean cell haemoglobin; MCHC = mean cell haemoglobin concentration; MCV = mean cell volume; NA = not applicable; ND = no data; PMN = polymorphonuclear neutrophil; PROM = premature rupture of membranes; RBC = red blood cell; ROC = receiver operating characteristic; ROM = rupture of membranes; SGA = small for gestational age; USA = United States of America; UTI = urinary tract infection; WCC = white cell count.

**Table 4.** Summary of geographical and economic distribution of included studies.

| WHO region | Country | Income classification | *n* studies |
| --- | --- | --- | --- |
| African Region (AFR) | Ethiopia | LIC | 1 |
|  | South Africa | UMIC | 2 |
|  | The Gambia | LIC | 1 |
|  | Zimbabwe | LMIC | 1 |
| Eastern Mediterranean Region (EMR) | Egypt | LMIC | 4 |
|  | Iran | LMIC | 2 |
|  | Jordan | LMIC | 1 |
|  | Pakistan | LMIC | 1 |
| European Region (EUR) | Bosnia and Herzegovina | UMIC | 1 |
|  | Russia | UMIC | 1 |
|  | Turkey | UMIC | 2 |
|  | Ukraine | LMIC | 1 |
| Region of the Americas (AMR) | Brazil | UMIC | 3 |
|  | Mexico | UMIC | 2 |
|  | Paraguay | UMIC | 1 |
| South-East Asian Region (SEAR) | Bangladesh | LMIC | 2 |
|  | India | LMIC | 37 |
|  | Indonesia | UMIC | 7 |
|  | Thailand | UMIC | 2 |
| Western Pacific Region (WPR) | China | UMIC | 9 |
|  | Papua New Guinea | LMIC | 1 |
|  | Philippines | LMIC | 3 |

One study conducted in multiple countries across multiple WHO regions and income classifications. LIC = low-income country; LMIC = lower middle-income country; UMIC = upper middle-income country; WHO = World Health Organization.

One study pooled data from both low-income and lower middle-income countries.^98^ Most studies were set in intensive care or special care admission units (*n=*64, 78%). The remainder included all live births at study sites (*n=*12, 15%), neonates presenting to emergency care services (*n=*3, 4%), all hospitalised neonates (*n*=1, 1%), or the setting was unclear (*n=*2, 2%).

In total, 24252 neonates were included across all studies. The median number of participants per study was 151 (range 36 to 3303, interquartile range [IQR] 200). Few studies restricted the study population based on gestational age or birthweight, with only 4 studies (5%) specifically investigating preterm neonates and 5 studies (6%) specifically investigating low or very low birthweight neonates. Most studies included neonates clinically suspected of sepsis or with specific maternal risk factors including chorioamnionitis (*n=*58, 71%).

Almost all studies included a positive blood and/or CSF culture in their outcome definition for sepsis (*n=*75, 91%). Of these, 18 (22% of all studies) also included clinical features or clinical suspicion of sepsis. One study used a consultant neonatologist’s clinical diagnosis of sepsis,^76^ one study used the International Classification of Diseases 10^th^ Revision criteria for sepsis,^77^ and in three studies the outcome was unclear.

### Model characteristics

The 82 included studies performed 109 evaluations validating 44 distinct models (Table 3).^22–25,32,33,40,46,47,49–51,54,56,57,63,68,72,76–78,81,83,86,87,90,92,98–101,103–113^ The most frequently validated model was the Hematological Scoring System by Rodwell et al. (*n=*32, 39% of studies; including studies that made minor modifications to the original model).^112^ Most models were only validated in one study (*n=*34, 77% of models).

A total of 135 predictors of sepsis were included across all models, of which 82 were clinical parameters (signs, symptoms or risk factors) and 53 were laboratory parameters (see supplementary appendix). The median number of predictors per model was 6 (range 2 to 110, IQR 4). 14 models (32%) included only clinical parameters, 12 models (27%) included only laboratory parameters, and 18 models (41%) included both. The commonest laboratory parameters were white cell count (*n=*17 models, 39%), C-reactive protein (CRP) (*n=*16 models, 36%) and platelet count (*n=*15 models, 34%). The commonest clinical parameters were neonatal fever (*n=*13 models, 30%) and gestational age (*n=*11 models, 25%).

Most models were developed using logistic regression (*n=*16 models, 36%) (often with stepwise selection to select predictors) or consisted of a scoring system based on univariable predictor performance or literature review and expert opinion (*n=*10 models, 23%).

### Model performance

Model performance was principally reported using sensitivity and/or specificity (with or without a confusion matrix); only 4 studies (5%) did not report either metric. 32 studies (39%) reported area under the receiver operating characteristic curve (AUC). Less frequent methods of quantifying performance included predictive values, likelihood ratios, accuracy, change in antibiotic use, and mortality statistics. Across all 109 evaluations, median sensitivity was 81% (range 3% to 100%) and median specificity was 83% (range 11% to 100%).

#### Performance stratified by clinical vs. laboratory parameters

In models containing both clinical and laboratory parameters (*n=*24 evaluations), median sensitivity was 73% (range 3% to 100%) and median specificity was 80% (range 18% to 98%). In models containing only clinical parameters (*n=*23 evaluations), median sensitivity was 67% (range 3% to 100%) and median specificity was 72% (range 11% to 99%). In models containing only laboratory parameters (*n=*62 evaluations), median sensitivity was 83% (range 9% to 100%) and median specificity was 86% (range 33% to 100%).

#### Performance stratified by target population

Models validated in a population with existing clinical suspicion of sepsis (due to signs and symptoms and/or presence of maternal risk factors for sepsis) had a median sensitivity of 82% (range 3% to 100%) and median specificity of 84% (range 18% to 100%) (*n=*75 evaluations). In comparison, models evaluated in the general population had a median sensitivity of 77% (range 15% to 100%) and median specificity of 80% (range 11% to 100%) (*n=*34 evaluations).

#### Performance stratified by sepsis timing

In models developed to diagnose both EOS and LOS (*n=*59 evaluations), median sensitivity was 82% (range 3% to 100%) and median specificity was 85% (range 33% to 100%). In models to diagnose only EOS, median sensitivity was 82% (range 9% to 100%) and median specificity was 82% (range 11% to 98%) (*n=*29 evaluations). In models to diagnose only LOS, median sensitivity was 57% (range 3% to 100%) and median specificity was 75% (range 18% to 99%) (*n=*21 evaluations).

## DISCUSSION

Our scoping review provides a comprehensive overview of CPMs to diagnose neonatal sepsis in LMICs. Previous reviews of CPMs to diagnose neonatal sepsis have been published, but we included more studies than were identified in existing reviews despite them additionally including studies from high-income countries.^11–14^ The breadth of literature highlights the need for, and academic interest in, effective risk stratification for neonatal sepsis in LMICs.

Several common themes emerged from our review. First, 99% of studies were conducted in middle-income countries, with only one study including neonates born in a low-income country,^98^ and no studies conducted exclusively in a low-income country. Furthermore, fewer studies were conducted in the African region than any other WHO region, despite the high burden of neonatal sepsis and slower progress in neonatal mortality in these countries.^114,115^ Two thirds of models required access to at least basic laboratory facilities. Access to laboratory facilities is limited in many low-resource settings or turnaround times are too long to usefully inform management.^116^

Second, although many CPMs have been validated in LMICs, 78% of studies were conducted in neonatal intensive care or special care units and only 29% were validated in a population of neonates that includes infants without existing suspicion of sepsis. A substantial benefit of CPMs for neonatal sepsis in LMICs lies in their ability to promote early, targeted antibiotic therapy in an undifferentiated population of neonates, reducing antibiotic overuse and the resultant antimicrobial resistance and adverse neonatal outcomes.^6^ Whilst diagnostic decision support in high-risk neonates is useful, models are needed that can be applied at the time of birth to facilitate the rapid antimicrobial therapy required to reduce morbidity and mortality from EOS.^7^

Third, description of model performance and clinical implications was highly variable between studies. Most studies only presented sensitivity and specificity at arbitrarily selected classification thresholds; just over one third of studies reported easily comparable global metrics such as AUC. Furthermore, few studies included practical discussion of how the proposed models could be integrated into routine neonatal care in LMICs. This includes how the probability or score can be calculated by healthcare workers (e.g. using a paper proforma or a computer-based system). Most models were developed using logistic regression, but authors often simplify the final model to present a scoring system where each predictor is assigned an integer score. This benefits interpretation and application but can negatively impact the resulting predictions, particularly if continuous predictors are categorised.^8^

One solution could be to implement models using app or web-based interfaces for clinicians such as the Neotree platform, which consists of an android app that also includes data visualisation, linkage and export capabilities.^117^ This would also aid implementation of more flexible classification methods, such as the artificial neural network proposed by Helguera-Repetto et al.^47^ Other considerations that are rarely discussed is how management decisions should be made based on model predictions, such as appropriate classification thresholds and how these can be incorporated into a wider sepsis management pathway within neonatal units. These oversights could be addressed if authors adhere to the TRIPOD checklist, which specifically includes ‘implications’ of the model as a checklist item.^21^

Finally, three quarters of models were only validated in one study in a LMIC, often only in the derivation cohort (internal validation). This may lead to overoptimistic performance results due to overfitting.^8^ Several authors caution against the current focus on developing new models, and advocate for further validation (including external validation) of promising existing models.^118^ Additionally, few studies included in our review assessed calibration performance of their models, which is an especially important consideration when models are intended for decision support.^119^

Several limitations of our review should be considered. First, we included only published studies. It may be that individual neonatal units or networks have developed their own clinical prediction models or tools that are in local use but have not been published and therefore would not have been identified by our search. Centres may use existing clinical prediction models without publishing performance data, particularly if studies show poor model performance. Second, there was significant heterogeneity in study populations, definitions of sepsis, and classification thresholds, which makes comparing model performance between studies particularly challenging. Despite the high incidence of neonatal sepsis globally, there is no internationally accepted consensus definition of neonatal sepsis.^120^ A systematic review of model performance in specific populations and for specific definitions of sepsis would help to address this. Given the vast number of model variations presented in included studies, and the often arbitrarily selected classification thresholds at which model performance is reported, it is impractical for a single review to be completely comprehensive and there is necessarily a degree of selective reporting in our review. Finally, we included only studies published in English or Spanish and were unable to obtain full texts for 36 potentially eligible studies despite contacting authors. The nature of scientific publishing in LMICs means that studies are not always published in journals indexed in major biomedical databases. We identified 13 of the 82 included studies outwith our primary database searches through citation analysis and reference lists only (especially for studies published in India), which raises the possibility of retrieval bias influencing our review.

## CONTRIBUTORS

FF and MH conceived the idea for this review. SRN developed the review protocol and all authors reviewed and approved it before publication. SRN and SS performed the literature searches and extracted the data. DM, HG, MZ, SRN and SS screened the records. FF, MH, GC, KLD and MCB supervised the review process and provided clinical expertise for the manuscript. MCB translated records from Spanish and provided statistical advice. All authors critically edited and approved the final manuscript.

## DATA SHARING

Extracted data and code used to synthesise results will be uploaded at publication. A list of all records identified through database searches is available on request.

## ACKNOWLEDGEMENTS

The authors would like to thank Heather Chesters (Deputy Librarian at the UCL Great Ormond Street Institute of Child Health Library) for her advice and guidance when developing the search strategy for this review. This review was conducted without specific funding support.

