## Supplementary Material for "Clinical prediction models to diagnose neonatal sepsis in low-income and middle-income countries: a scoping review"

**SUPPLEMENTARY APPENDIX**

**Appendix 1.** Preferred Reporting Items for Systematic reviews and Meta-Analyses extension for Scoping Reviews (PRISMA-ScR) Checklist.

**Appendix 2.** Search strategies for each database.

**Appendix 3.** Data extraction form.

**Appendix 4.** Additional results.

**Supplementary File 1.** Full text decisions and exclusion reasons (.csv).

**Supplementary File 2.** Data extracted from included studies (.xlsx).

**Supplementary File 3.** Code used to analyse extracted data (.R).

**Appendix 1.** Preferred Reporting Items for Systematic reviews and Meta-Analyses extension for Scoping Reviews (PRISMA-ScR) Checklist.

| SECTION | ITEM | PRISMA-ScR CHECKLIST ITEM | REPORTED ON PAGE # |
|---|---|---|---|
| **TITLE** | | | |
| Title | 1 | Identify the report as a scoping review. | 1 |
| **ABSTRACT** | | | |
| Structured summary | 2 | Provide a structured summary that includes (as applicable): background, objectives, eligibility criteria, sources of evidence, charting methods, results, and conclusions that relate to the review questions and objectives. | 1 |
| **INTRODUCTION** | | | |
| Rationale | 3 | Describe the rationale for the review in the context of what is already known. Explain why the review questions/objectives lend themselves to a scoping review approach. | 2 |
| Objectives | 4 | Provide an explicit statement of the questions and objectives being addressed with reference to their key elements (e.g., population or participants, concepts, and context) or other relevant key elements used to conceptualize the review questions and/or objectives. | 2 |
| **METHODS** | | | |
| Protocol and registration | 5 | Indicate whether a review protocol exists; state if and where it can be accessed (e.g., a Web address); and if available, provide registration information, including the registration number. | 3 |
| Eligibility criteria | 6 | Specify characteristics of the sources of evidence used as eligibility criteria (e.g., years considered, language, and publication status), and provide a rationale. | Table 1 |
| Information sources | 7 | Describe all information sources in the search (e.g., databases with dates of coverage and contact with authors to identify additional sources), as well as the date the most recent search was executed. | 3 |
| Search | 8 | Present the full electronic search strategy for at least 1 database, including any limits used, such that it could be repeated. | Appendix 2 |
| Selection of sources of evidence | 9 | State the process for selecting sources of evidence (i.e., screening and eligibility) included in the scoping review. | 3 |
| Data charting process | 10 | Describe the methods of charting data from the included sources of evidence (e.g., calibrated forms or forms that have been tested by the team before their use, and whether data charting was done independently or in duplicate) and any processes for obtaining and confirming data from investigators. | 3 |

| | | | |
|---|---|---|---|
| Data items | 11 | List and define all variables for which data were sought and any assumptions and simplifications made. | Appendix 3 |
| Critical appraisal of individual sources of evidence | 12 | If done, provide a rationale for conducting a critical appraisal of included sources of evidence; describe the methods used and how this information was used in any data synthesis (if appropriate). | Not applicable |
| Synthesis of results | 13 | Describe the methods of handling and summarizing the data that were charted. | 3 |
| **RESULTS** | | | |
| Selection of sources of evidence | 14 | Give numbers of sources of evidence screened, assessed for eligibility, and included in the review, with reasons for exclusions at each stage, ideally using a flow diagram. | Figure 1 |
| Characteristics of sources of evidence | 15 | For each source of evidence, present characteristics for which data were charted and provide the citations. | 3-4 |
| Critical appraisal within sources of evidence | 16 | If done, present data on critical appraisal of included sources of evidence (see item 12). | Not applicable |
| Results of individual sources of evidence | 17 | For each included source of evidence, present the relevant data that were charted that relate to the review questions and objectives. | Table 2 and 3 |
| Synthesis of results | 18 | Summarize and/or present the charting results as they relate to the review questions and objectives. | 4-5 |
| **DISCUSSION** | | | |
| Summary of evidence | 19 | Summarize the main results (including an overview of concepts, themes, and types of evidence available), link to the review questions and objectives, and consider the relevance to key groups. | 5-6 |
| Limitations | 20 | Discuss the limitations of the scoping review process. | 6 |
| Conclusions | 21 | Provide a general interpretation of the results with respect to the review questions and objectives, as well as potential implications and/or next steps. | 5-6 |
| **FUNDING** | | | |
| Funding | 22 | Describe sources of funding for the included sources of evidence, as well as sources of funding for the scoping review. Describe the role of the funders of the scoping review. | 7 |

Adapted from: Tricco AC, Lillie E, Zarin W, O'Brien KK, Colquhoun H, Levac D, et al. PRISMA Extension for Scoping Reviews (PRISMAScR): Checklist and Explanation. Ann Intern Med. 2018;169:467–473. doi: 10.7326/M18-0850.

**Appendix 2.** Search strategies for each database.

**Ovid MEDLINE**

*Ovid MEDLINE(R) and Epub Ahead of Print, In-Process & Other Non-Indexed Citations and Daily*

| | |
|---|---|
| 1 | exp Infant, Newborn/ |
| 2 | (neonat* or newborn* or new born* or baby or babies or premature or preterm or infant* or low birth weight or LBW or VLBW or ELBW or NICU*).ti,ab,kw. |
| 3 | 1 or 2 |
| 4 | exp Sepsis/ |
| 5 | (sepsis or septic* or bacter?emia).ti,ab,kw. |
| 6 | 4 or 5 |
| 7 | Decision Support Techniques/ or Neonatal Screening/ |
| 8 | ((predict* or diagnos* or screen* or identif* or manag*) adj5 (model* or rule* or scor* or tool* or algorithm* or decision tree* or pathway* or calculator*)).ti,ab,kw. |
| 9 | 7 or 8 |
| 10 | 3 and 6 and 9 |

**Ovid Embase**

| | |
|---|---|
| 1 | Newborn/ |
| 2 | (neonat* or newborn* or new born* or baby or babies or premature or preterm or infant* or low birth weight or LBW or VLBW or ELBW or NICU*).ti,ab,kw. |
| 3 | 1 or 2 |
| 4 | exp sepsis/ |
| 5 | (sepsis or septic* or bacter?emia).ti,ab,kw. |
| 6 | 4 or 5 |
| 7 | exp decision support system/ or newborn screening/ |
| 8 | ((predict* or diagnos* or screen* or identif* or manag*) adj5 (model* or rule* or scor* or tool* or algorithm* or decision tree* or pathway* or calculator*)).ti,ab,kw. |
| 9 | 7 or 8 |
| 10 | 3 and 6 and 9 |

**Scopus**

TITLE-ABS-KEY((neonat* OR newborn* OR "new born*" OR baby OR babies OR premature OR preterm OR infant* OR "low birth weight" OR lbw OR vlbw OR elbw OR nicu*) AND (sepsis OR septic* OR bacter?emia) AND ((predict* OR diagnos* OR screen* OR identif* or manag*) W/5 (model* OR rule* OR scor* OR tool* OR algorithm* OR "decision tree*" OR pathway* OR calculator*)))

**Web of Science**
*Core Collection*

TS=((neonat* OR newborn* OR "new born*" OR baby OR babies OR premature OR preterm OR infant* OR "low birth weight" OR lbw OR vlbw OR elbw OR nicu*) AND (sepsis OR septic* OR bacter$emia) AND ((predict* OR diagnos* OR screen* OR identif* or manag*) NEAR/5 (model* OR rule* OR scor* OR tool* OR algorithm* OR "decision tree*" OR pathway* OR calculator*)))

**Global Index Medicus**
*All indexes: LILCAS (Americas), WPRIM (Western Pacific), IMSEAR (South-East Asia), IMEMR (Eastern Mediterranean), AIM (Africa)*

((mh:("Infant, Newborn")) OR (tw:(neonat* OR newborn* OR "new born*" OR baby OR babies OR premature OR preterm OR infant* OR "low birth weight" OR lbw OR vlbw OR elbw OR nicu*))) AND ((mh:(“Sepsis”)) OR (tw:(sepsis OR septic* OR bacter*emia))) AND ((mh:(“Decision Support Systems, Clinical” OR “Neonatal Screening”)) OR (tw:(model* OR rule* OR scor* OR tool* OR algorithm* OR "decision tree*" OR pathway* OR calculator*)))

**Cochrane Library**
*Cochrane Database of Systematic reviews and Cochrane Central Register of Controlled Trials*

| | |
|---|---|
| 1 | MeSH descriptor: [Infant, Newborn] explode all trees |
| 2 | (neonat* or newborn* or “new born*” or baby or babies or premature or preterm or infant* or “low birth weight” or LBW or VLBW or ELBW or NICU*):ti,ab,kw |
| 3 | #1or#2 |
| 4 | MeSH descriptor: [Sepsis] explode all trees |
| 5 | (sepsis or septic* or bacter*emia):ti,ab,kw |
| 6 | #4or#5 |
| 7 | MeSH descriptor: [Decision Support Systems, Clinical] explode all trees |
| 8 | MeSH descriptor: [Neonatal Screening] explode all trees |
| 9 | ((predict* or diagnos* or screen* or identif* or manag* or estimat*) NEAR/5 (model* or rule* or scor* or tool* or algorithm* or “decision tree*” or pathway* or calculator*)):ti,ab,kw |
| 10 | #7 or #8 or #9 |
| 11 | #3 and #6 and #10 |

**Appendix 3.** Data extraction form.

| | |
|---|---|
| Study characteristics | |
| Study ID | Title, author, year |
| Country | |
| Context | E.g. NICU/SCBU admissions, all babies born at study site, presenting to emergency care services |
| Number of included participants | |
| Characteristics of included participants | E.g. preterm, low birthweight |
| Objectives | |
| Outcome definition | E.g. blood culture, CSF culture, clinical diagnosis |
| Study results | |
| Name of model | |
| Modelling methods | E.g. logistic regression, scoring system |
| Predictors in final model | Signs, symptoms, risk factors, laboratory parameters |
| Model performance | Sensitivity, specificity, predictive values, likelihood ratios, AUC, accuracy, acceptability, antibiotic use, mortality, any other factors |

**Appendix 4.** Additional results.

Included studies by World Health Organization (WHO) region.

| **WHO region** | ***n* studies** |
|---|---|
| African Region (AFR) | 4 |
| Eastern Mediterranean Region (EMR) | 8 |
| European Region (EUR) | 5 |
| Region of the Americas (AMR) | 6 |
| South-East Asian Region (SEAR) | 48 |
| Western Pacific Region (WPR) | 12 |

One study conducted in multiple WHO regions.

Included studies by World Bank 2020 income classification.

| **Income classification** | ***n* studies** |
|---|---|
| Lower income | 1 |
| Lower middle-income | 52 |
| Upper middle-income | 30 |

One study conducted in countries from multiple income classifications.

Predictors in included models.

| **Predictor** | ***n* models** |
|---|---|
| Clinical features | |
| Abdominal distension | 4 |
| Abnormal heart rate | 8 |
| Abnormal temperature | 3 |
| Age at admission | 3 |
| Albumin use | 1 |
| Altered behaviour, altered consciousness or lethargy | 9 |
| Apgar-1 | 3 |
| Apgar-5 | 3 |
| Apnoea | 2 |
| Asphyxia | 1 |

| | |
|---|---|
| Birthweight | 7 |
| Bulging fontanelle | 1 |
| Cardiovascular symptoms | 1 |
| Catheter | 1 |
| Cervicovaginitis | 1 |
| Chorioamnionitis | 4 |
| Central nervous system symptoms | 1 |
| Colour or pallor | 2 |
| Continuous positive airway pressure | 1 |
| Cough | 1 |
| Dehydration | 1 |
| Diastolic blood pressure | 1 |
| End-diastolic flow | 1 |
| Equivocal appearance | 1 |
| Evidence of focal infection | 1 |
| Feed intolerance or increased aspirates | 2 |
| Feeding difficulty | 4 |
| Fetal or neonatal morbidity | 4 |
| Foul liquor | 3 |
| Fungal infection | 1 |
| Gastrointestinal symptoms | 1 |
| Gestational age | 11 |
| Hospital length of stay | 1 |
| Hypotension | 3 |
| Hypothermia | 2 |
| Hypoxia or oxygen saturation | 3 |
| ICU length of stay | 1 |
| Ill appearance | 3 |
| In-hospital mortality | 1 |
| Increased oxygen or ventilation requirement | 4 |
| Infective symptoms | 1 |

| | |
|---|---|
| Inotropes or dopamine or vasoactive drug use | 3 |
| Intrapartum antibiotics | 1 |
| Intrauterine growth restriction | 1 |
| Intubated or mechanical ventilation | 3 |
| Labour >24h | 1 |
| Length of stay pre-sepsis | 1 |
| Local incidence of sepsis | 1 |
| Maternal age | 3 |
| Maternal Group B streptococcus | 1 |
| Maternal history of abortion | 1 |
| Maternal morbidity | 2 |
| Maternal parity or gravidity | 1 |
| Maternal temperature or fever | 2 |
| Neonatal fever | 13 |
| Nitrous oxide | 1 |
| Perfusion or capillary refill time | 4 |
| Premature rupture of membrane | 1 |
| Prenatal glucocorticoid use | 2 |
| Previous hospitalisation or illness | 2 |
| Previously healthy | 3 |
| Received antibiotics | 4 |
| Received recent immunisation | 1 |
| Respiratory insufficiency or distress | 7 |
| Respiratory rate | 4 |
| Respiratory symptoms | 1 |
| Rupture of membranes duration | 6 |
| Season | 1 |
| Seizures | 1 |
| Sex | 5 |
| Signs of encephalopathy or abnormal neurological examination | 1 |

| | |
|---|---|
| Skin symptoms | 1 |
| Small for gestational age | 1 |
| Systolic blood pressure | 1 |
| Total parenteral nutrition | 2 |
| Type of birth | 2 |
| Urinary tract infection | 1 |
| Umbilical cord winding | 1 |
| Umbilical or central venous catheter | 6 |
| Unclean vaginal examination | 1 |
| Vomiting | 1 |
| Well appearance | 5 |
| Laboratory parameters | |
| Calcium | 1 |
| CD34 | 1 |
| CD69 | 1 |
| CD95 | 1 |
| Cell volume | 1 |
| Chest x-ray | 4 |
| Conductivity for internal composition of cell | 1 |
| Creatinine | 1 |
| CRP | 16 |
| CSF white cell count | 4 |
| Degenerative changes in polymorphonuclear cells | 2 |
| Delta neutrophil index | 1 |
| Eosinophil count or percentage | 1 |
| Gastric acid cytology | 1 |
| Glucose | 3 |
| Haemoglobin | 1 |
| Haematocrit | 1 |
| I:M ratio | 2 |
| I:T ratio | 9 |

| | |
|---|---|
| IL-6 | 2 |
| IL-27 | 1 |
| Immature polymorphonuclear cell count or percentage, band cell count or percentage, or neutrophil bandaemia | 4 |
| Lactate | 1 |
| Light scatter for cytoplasmic granularity and nuclear structure | 1 |
| Lower median angle light scatter | 1 |
| Lymphocyte count or percentage | 2 |
| Lymphocyte:CRP ratio | 1 |
| Lymphocytes with expression AnnexinV-FITC+PI | 1 |
| Mean cell haemoglobin | 1 |
| Mean cell haemoglobin concentration | 1 |
| Mean cell volume | 1 |
| Mean neutrophil volume | 1 |
| Mean platelet volume | 1 |
| Micro ESR or ESR | 3 |
| Monocyte count or percentage | 2 |
| Monocyte:lymphocyte ratio | 1 |
| Neutrophil count or percentage | 11 |
| Neutrophil:lymphocyte ratio | 1 |
| Nitric oxide | 1 |
| Partial thromboplastin time | 1 |
| pH | 2 |
| Platelet count | 15 |
| Platelet distribution width | 1 |
| Platelet:lymphocyte ratio | 1 |
| Positive microbiological test | 1 |
| Procalcitonin | 4 |
| Prothrombin time | 1 |
| Red blood cell count | 1 |
| Stool white cell count | 3 |

| | |
|---|---|
| Thyroid function | 1 |
| Total polymorphonuclear count | 1 |
| Urine white cell count | 4 |
| Visual placental changes | 1 |
| White cell count | 17 |
